# Novel point-of-care ultrasound (POCUS) technique to modernize the JVP exam and rule out elevated right atrial pressures

**DOI:** 10.1101/2021.10.14.21264891

**Authors:** Larry Istrail, Maria Stepanova

**Author notes:** Correspondence to: Larry Istrail.

## Abstract

Accurate assessment of the jugular venous pressure (JVP) and right atrial pressure (RAP) has relied on the same bedside examination method since 1930. While this technique provides a rough estimate of right sided pressures, it is limited by poor sensitivity and overall diagnostic inaccuracy. The internal jugular vein (IJV) is difficult to visualize in many patients and relies on an incorrect assumption that the right atrium lies 5 centimeters below the sternum. Point-of-care ultrasound (POCUS) offers an alternative method for more precisely estimating JVP and RAP. We propose a novel method of measuring the right atrial depth (RAD) using a sonographic measurement of the depth of the posterior left ventricular outflow tract as a surrogate landmark to the center of the right atrium when viewed in the parasternal long axis view. This is combined with determination if JVD was present at the supraclavicular point. Sensitivity, specificity, PPV, NPV of JVD at the supraclavicular point was 70%, 76%, 59%, 91% respectively. These values were confounded by the lack of standardization of zero reference landmarks (ZRLs) used during the right heart catheterizations. When the RAD measurement was adjusted to account for measurement error the sensitivity of JVD at supraclavicular point for elevated RAP improved to 90% with negative predictive value of 96%. This may offer a rapid and reliable method for ruling out elevated RAP and increase objectivity in our volume status assessment.

## Background

The evaluation of neck veins to assess for elevated pressures dates back to Sir Thomas Lewis in 1930.^1^ It involves visually identifying the internal jugular venous pulsations on the lateral neck and then measuring the distance from this point down to the sternum. This distance in centimeters is then added to the estimated distance from the sternum down to the center of the right atrium, classically assumed to be 5 centimeters.^2^ This JVP is viewed as a surrogate for right atrial pressure (RAP) which is itself used as a surrogate for pulmonary capillary wedge pressure and overall volume status, making accurate JVP estimation crucial to our clinical assessment. However, there are many limitations to the current method that result in poor sensitivity and low overall diagnostic accuracy.^2–8^

The first is the lack of certainty offered by visual inspection of the neck veins to determine if the jugular venous pulsation is present above the clavicle or not. The jugular venous pulsation may not be detected for multiple reasons including: 1) the jugular pulsations are obscured below the clavicle due to normal or low JVP, 2) The jugular pulsations reside above the mandible due to very elevated JVP, 3) Patient’s body habitus or neck girth prevents visualization, or 4) clinician error.

The other major limitation to our classic JVP exam is the assumption that the right atrium lies 5 centimeters below the sternum. This originated from a 1946 study in which this right atrial depth (RAD) was estimated from a supine chest x-ray.^9^ More recent studies based on CT scan measurements suggest the RAD is in fact quite variable depending on the patient’s age, smoking status, AP diameter of the chest, and the angle of the head of the bed. Seth et al found the median distance from the anterior chest wall to the center of the right atrium to be 8.5 centimeters at 30 degrees and 9.9 centimeters are 45 degrees, with individual variation from 5 to 15 centimeters.^10^ Kovacs et al found the distance from the anterior thoracic surface to the mid right atrium to be 9 centimeters on average, with the right atrium located 5 centimeters below the sternum only 29% of the time.^11^ Ultimately these limitations result in clinical uncertainty and the need for invasive right heart catheterization (RHC).

Point-of-care ultrasound (POCUS) has the potential to overcome these limitations. It has proven to be an accurate way to visualize the IJV and detect the meniscus, even in cases when visual inspection fails.^12–14^ Two studies have attempted to estimate JVP and RAP using POCUS, both underestimating the actual RAP.^12,15^ We hypothesize this is due to the reliance on the 5 cm RAD assumption, and that for accurate estimation of JVP, the RAD must be measured in each patient. This was evidenced by Xing et al^16^ that showed a near-perfect correlation between ultrasound estimation and CVP. However, their methods were complex and not applicable to a physical exam at the patient’s bedside. We have developed a simple and novel method for estimating RAD that may allow for more accurate JVP and RAP estimation during the physical examination.

## Methods

Patients undergoing RHCs were recruited for this study if they were 21 years old or over and required RHC for any purpose. Patients were excluded from the study based on the following: 1) Right internal jugular vein catheter present; 2) intubated; 3) congenital heart disease history.

### Ultrasound Examination

Within 2 hours before or after the RHC, the patient’s head of the bed was placed at 45 degrees. A Butterfly IQ+ probe was placed along the left sternal border and a cardiac parasternal long axis view was acquired. The probe was positioned perpendicularly to the chest wall in order to measure the distance from the chest wall down to where the non-coronary cusp of the aortic valve attaches to the posterior wall of the left ventricular outflow tract (LVOT) (Figure 1).

**Figure 1:**
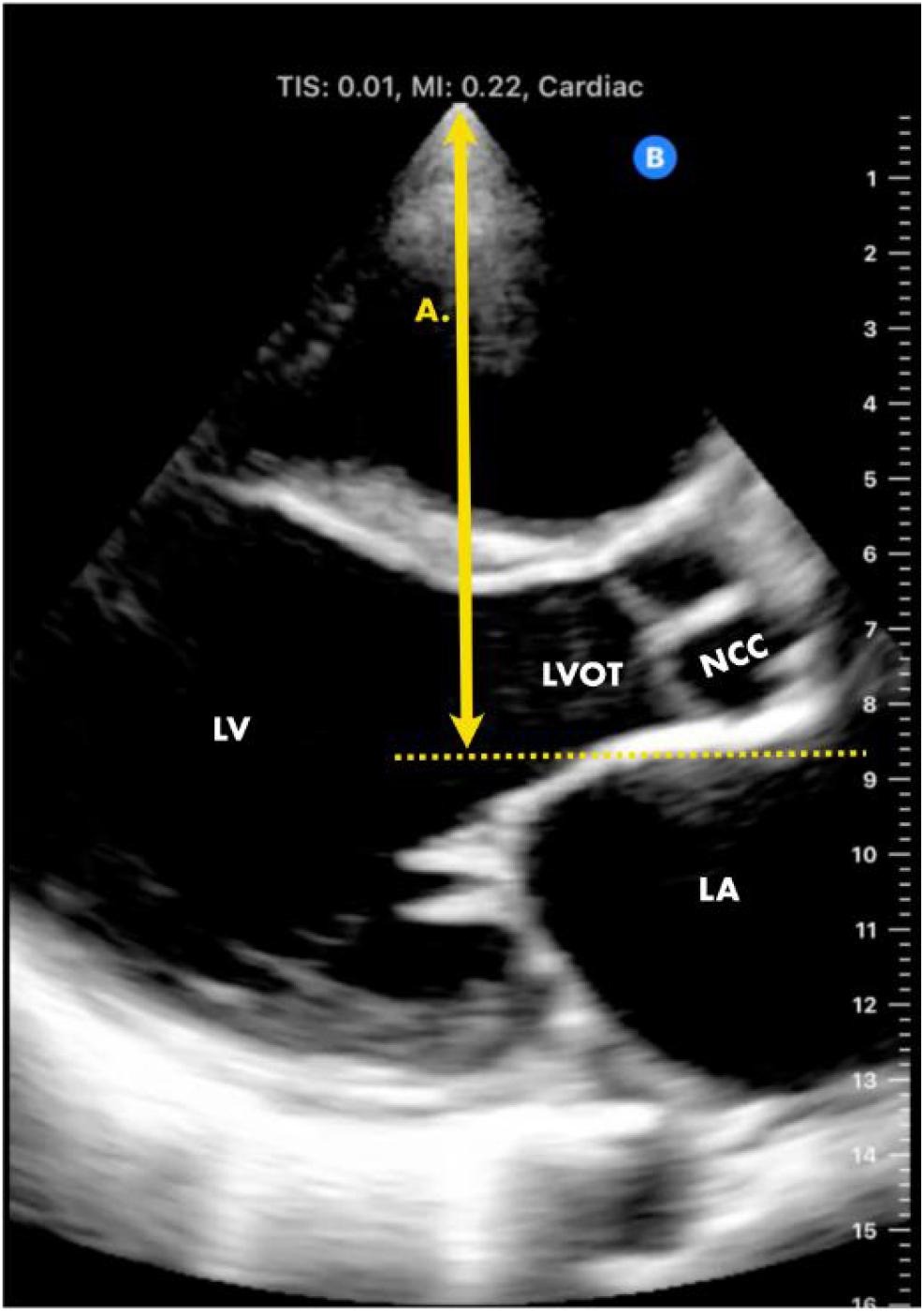
Right Atrial Depth (RAD) A. Right atrial depth (RAD). With the probe perpendicular to the chest wall, the distance from the chest wall to the posterior wall of the LVOT where the non-coronary cusp (NCC) attaches is measured.

This landmark was chosen as a surrogate because it sits in the same approximate coronal plane as the center of the right atrium and is easily detectable with a patient supine. This distance was recorded as the RAD.

The probe was then gently placed just superior to the clavicle at the supraclavicular point. In the transverse view, the IJV was visualized. A clip of the IJV at this point was recorded. IJV shape, degree of collapsibility, or presence of jugular venous distention (JVD) was determined. JVD was considered present at the supraclavicular point if the vein was distended with minimal or absent jugular venous pulsations. If the vein was distended but collapsed completely, or near completely, then JVD was deemed not present.

The estimated RAP value was calculated in two discrete ways depending on if there was JVD present at the supraclavicular point or not. If JVD was present at the supraclavicular point, then with the ultrasound probe remaining in the transverse plane, it was slid cranially to identify the jugular venous collapse point where the venous walls collapse completely. The probe was then rotated 90 degrees into the longitudinal plane to confirm this point. This is the meniscus of the blood column that corresponds to the jugular venous pulse (Figure 2). We will call it the ‘Wine Bottle Sign,’ due to the previous description of its resemblance to the top of a wine bottle in the longitudinal plane.^17^ The vertical distance from this point down to the sternum was then measured in centimeters using a ruler. This value was then added to the RAD in cm to estimate RAP.

**Figure 2:**
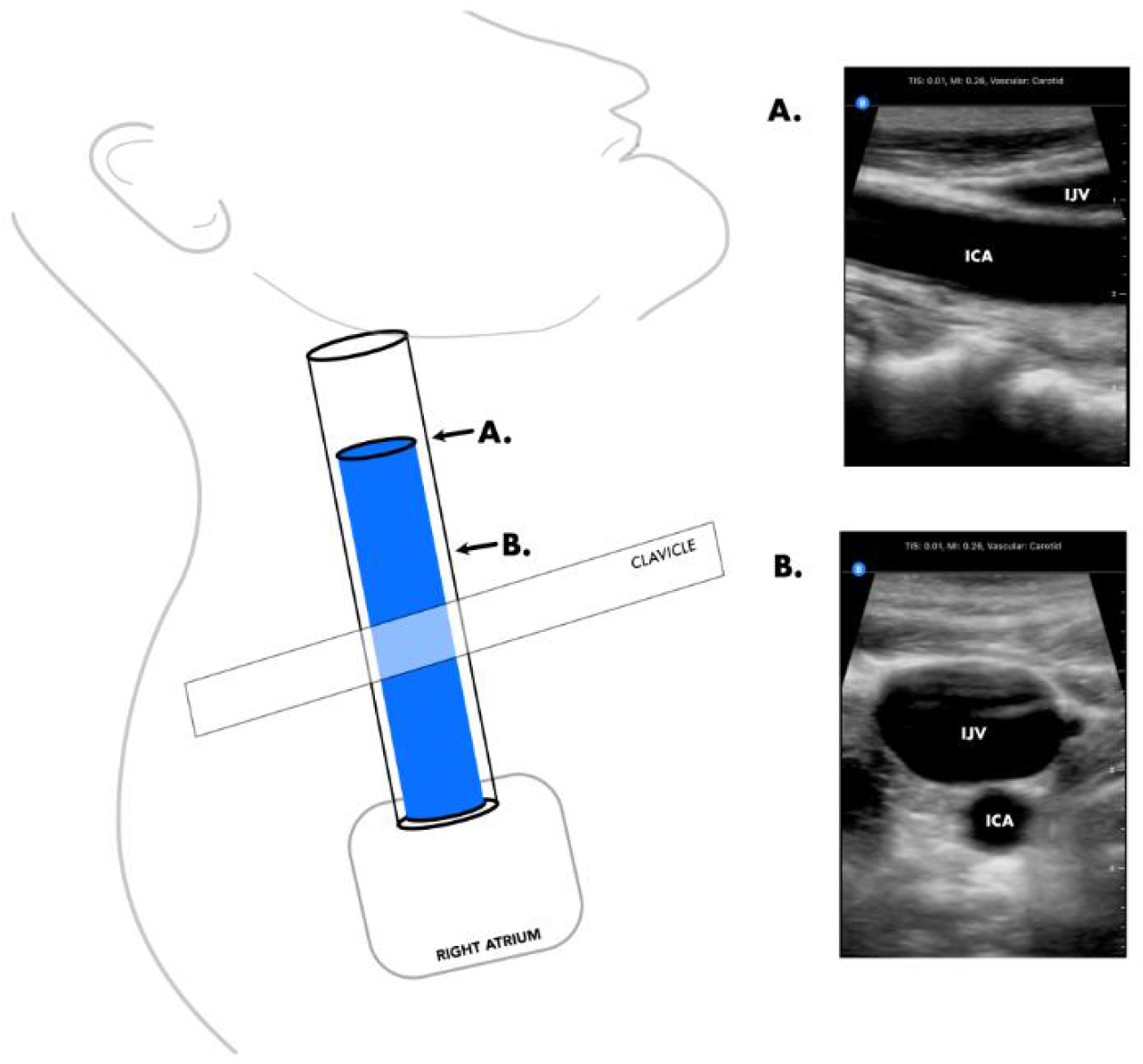
Appearance of Jugular Venous Distention and wine Bottle Sign with POCUS. **A:** wine Bottle Sign: Meniscus of blood column in internal jugular vein (IJV) where juglar venous pulsations most prominent. IJV assumes a triangular appearance at the meniscus. IJV lies just superficial to internal carotid artery (ICA). **B:** Presence of JVD at supraclavicular point. IJV is dilated with minimal of absent pulsations

If JVD was absent at the supraclavicular point and the vein was completely collapsing, then jugular venous pulsations were deemed present and JVD absent. In order to estimate the pressure in this case, the ultrasound probe remained at the supraclavicular point, maintaining a transverse view of the IJV. The head of the bed was then lowered to 30 degrees. The IJV was assessed for partial or full engorgement. If the vein does not engorge to a greater degree than at the 45-degree position, then the head of the bed is dropped further to zero degrees, and the presence of partial or full engorgement was recorded. If the vein engorged at 30 degrees, then the estimated RAP in cm H_2_O was RAD x 0.75. If the IJV engorged at zero degrees, then the RAP was estimated to be RAD x 0.5. If the vein still did not engorge at zero degrees, then the RAP was estimated to be RAD x 0.25. The estimated pressure in cm H_2_O was then converted to mmHg by multiplying by 0.735.

### Study Outcomes

The study included a qualitative and quantitative estimate of RAP. The primary aim of the qualitative phase was to determine if JVD at the supraclavicular point would predict elevated right atrial pressures greater than or equal to the RAD. The aims of the quantitative phase were to calculate an estimated RAP and to assess its association with the actual RAP from RHC.

### Statistical Analysis

The results of the qualitative phase were summarized as a 2×2 table using RAD as a reference for actual RAP; that table was further used to yield sensitivity (proportion of patients with RAP ≥ RAD among those with JVD at supraclavicular point), specificity (proportion of patients with RAP < RAD among those without JVD), positive predictive value (proportion of patients with JVD at supraclavicular point among those with RAP ≥ RAD), and negative predictive value (proportion of patients without JVD at supraclavicular point among those with RAP < RAD). We also ran a round of sensitivity analysis using the RAD minus 1 mmHg instead of the original RAD value, to account for variation in RHC zeroing techniques and potential inaccuracy of this cutoff measurement. That way, patients with actual RAP measured less than the original RAD by a value of no more than 1 mmHg would also be considered as predicted high RAP.

The outcome of the quantitative phase was high RAP defined as actual RAP ≥ 9 mmHg.^18^ The overall accuracy of prediction of high RAP by estimated RAP was assessed using the area under the ROC curve (AUC). Wilcoxon non-parametric test was used to compare estimated RAP values between patients with and without high RAP. Estimated RAP was also summarized by the tertiles of actual RAP distribution using median and inter-quartile range (IQR), and correlation of actual RAP with estimated RAP was calculated.

All analyses were run using SAS 9.4 (SAS Institute, Cary, NC, USA). All patients provided informed consent for participation in the study. The study was approved by the Inova Institutional Review Board. All measurements were taken and entered into the research database while blinded to the results of the RHC. After the RHC was completed, the actual RAP, RV systolic pressure, RV diastolic pressure, pulmonary capillary wedge pressure, and pulmonary artery pressures were recorded.

## Results

Fifty-five patients were prospectively enrolled, but two were later excluded due to the inability to visualize the posterior LVOT in the parasternal long axis view due to the presence of breast implants. In addition, two patients were scheduled for both left and right heart catheterization but underwent left heart catheterization only. Therefore, 51 patients were included in the final analysis.

Patient characteristics are summarized in Table 1. The average age was 68 years. The sample was 63% male, 37% female. The mean BMI was 29.5. The average right atrial depth was 10.16 cm with a range from 8.0-13.1 cm. Overall, 57% (29 subjects) had RAP > 5mmHg and 27% (14 subjects) had RAP ≥ 10 mmHg. In addition, 56% had a normal ejection fraction, 35% had a reduced ejection fraction, 6% had a severely reduced ejection fraction.

**TABLE 1:** Baseline Characteristics.

|  | High RAP: JVD at supraclavicular point | Normal RAP: No JVD at supraclavicular point | p-value | All |
| --- | --- | --- | --- | --- |
| **N** | **10** | **41** |  | **51** |
| Age, years | 77.1 +/- 14.1 | 66.1 +/- 11.0 | 0.0042 | 68.2 +/- 12.3 |
| Male | 4 (40.0%) | 28 (68.3%) | 0.10 | 32 (62.7%) |
| BMI | 28.8 +/- 5.0 | 29.7 +/- 6.2 | 0.78 | 29.5 +/- 6.0 |
| Normal EF | 3 (33.3%) | 24 (61.5%) | 0.12 | 27 (56.3%) |
| Hyperdynamic EF | 0 (0.0%) | 1 (2.6%) | 0.63 | 1 (2.1%) |
| Reduced EF | 6 (66.7%) | 11 (28.2%) | 0.0297 | 17 (35.4%) |
| Severely reduced EF | 0 (0.0%) | 3 (7.7%) | 0.39 | 3 (6.3%) |
| Predicted high RAP (Actual RAP ≥ RAD) | 7 (70.0%) | 10 (24.4%) | 0.0061 | 17 (33.3%) |
| RAD, cm | 9.9 +/- 1.13 | 10.24 +/- 1.25 | 0.64 | 10.16 +/- 1.22 |
| Actual RAP, mmHg | 12.5 +/- 7.5 | 5.88 +/- 3.14 | 0.0021 | 7.18 +/- 5.01 |
| Estimate RAP, mmHg | 11.7 +/- 2.9 | 4.21 +/- 1.09 | <0.0001 | 5.68 +/- 3.39 |

In the qualitative phase of the study, we found that 10/51 patients had a vein distention at supraclavicular point (Table 2). Of those, 7 had actual RAP measured above the RAD, resulting in the sensitivity value of 70%, accompanied by specificity of 76%, PPV 59%, NPV 91% (Table 2). However, of the three patients who had JVD at the supraclavicular point but RAP was less than RAD, two had it within the cutoff error margin of 1 mmHg (8.1 mmHg vs. 8 mmHg, 7.4 mmHg vs. 7 mmHg). Therefore, in a round of sensitivity analysis with a modified RAD, sensitivity increased to 90% with NPV of 96% (Table 2).

**Table 2:**
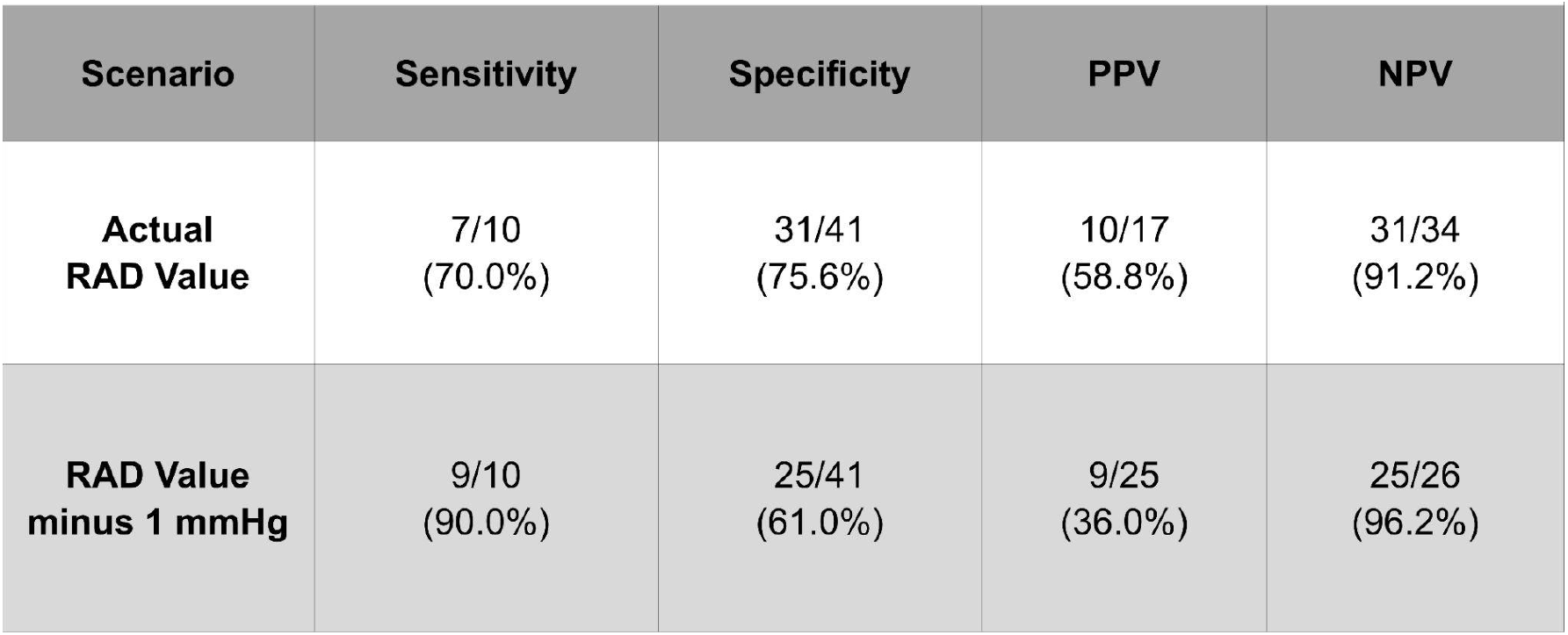
High RAP vs. prediction of high RAP.

| Scenario | Sensitivity | Specificity | PPV | NPV |
| --- | --- | --- | --- | --- |
| **Actual**<br>**RAD Value** | 7/10<br>(70.0%) | 31/41<br>(75.6%) | 10/17<br>(58.8%) | 31/34<br>(91.2%) |
| **RAD Value**<br>**minus 1 mmHg** | 9/10<br>(90.0%) | 25/41<br>(61.0%) | 9/25<br>(36.0%) | 25/26<br>(96.2%) |

For the quantitative phase of the study, the distribution of estimated RAP values by the tertiles of actual RAP is shown in Figure 3. The correlation of estimated RAP with actual RAP was +0.68 (p<0.0001). The difference in estimated RAP values between patients with and without high actual RAP (≥9 mmHg) was significant: mean (SD) 7.9 (4.3) vs. 4.7 (2.4), p=0.0041. Finally, RAP estimate was predictive of high actual RAP with AUC of 0.76 (p<0.0001) while the Youden’s criterion (estimated RAP > 4 mmHg) returned sensitivity of 87%, specificity 56%, PPV 45%, and NPV 91%.

**Figure 3:**
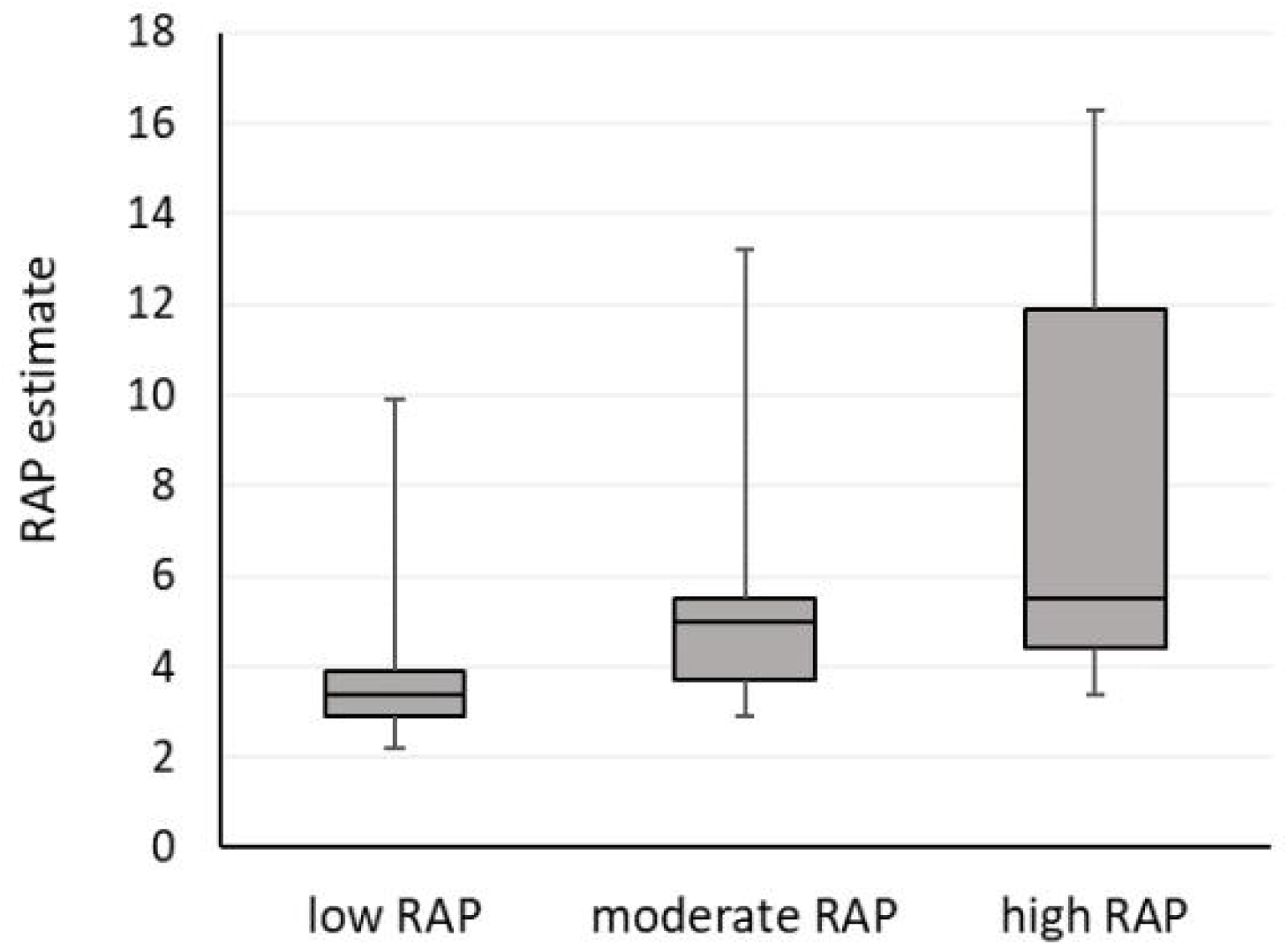
Distribution of RAP. Estimated RAP (media (IQR)) by tertiles of actual RAP (low RAP ≤ 4 mmHg, moderate RAP > 4 to <9 mmHg, high RAP ≥ 9 mmHg)

## Discussion

The main findings of this study are that a simple bedside ultrasound technique has the potential to effectively rule out elevated RAP regardless of a patient’s body habitus. Clinically this information is essential at the bedside for determining a need for diuretics, especially in patients whose clinical exam is limited by their neck size, BMI, or other factors.

Other studies have already shown that visualizing the jugular vein with ultrasound is reliable and feasible in every patient; however, these studies still rely on the 5-centimeter assumption for right atrial depth which may explain their underestimation of actual RAP. This is likely due to the fact that the RAD is on average much deeper than 5 centimeters and may change with the angle of the head of the bed. In our study - using the non-coronary cusp attachment to the posterior LVOT as a proxy, the average distance to the right atrium at the 45-degree position was 10.16 cm, which is similar to the 9 centimeters in Kovacs et al^11^ and 9.9 cm at 45 degrees in Seth et al^10^ which both relied on measurements from CT. Accurate assessment of RAP within 0.2 cm has already been demonstrated with ultrasound when the right atrial depth is measured in each patient. Though the method used by Xang et al^16^ is not practical for a physical exam, this strategy has been shown to be extremely accurate.

This study also gleaned insight into the difficulties of using a RHC as the gold standard measurement since it is subject to its own variation. There are various proposed zero reference levels (ZRLs) used to zero the catheter at the start of a RHC: 5 centimeters below the sternum, ⅓ the thoracic diameter, mid-thoracic level, and 10 cm above the table.^11^ The resulting pressure measurements can be significantly altered depending on which one is chosen. Using CT scan measurements, Kovacs et al showed that 98% of ZRLs at ⅓ the thoracic diameter were located somewhere in the right atrium, while only 21% of those at 5 centimeters below the sternal angle were, and only 3.6% of ZRLs 10 cm above the table were. This could change the resulting pressure by as much as 15.4 mmHg in the obese patients included in this study. Different ZRLs would have changed the diagnosis of pulmonary hypertension in 21% of cases and led to a change of pulmonary arterial wedge pressure class in 31% of cases. Similarly, when the catheter is zeroed in the cath lab, it is often performed once the patient is fully draped making the precise identification of the ZRL more difficult. It is therefore challenging to compare the accuracy of this new method using the posterior wall of the LVOT as a landmark without controlling for which ZRL was used. This likely caused some of the discrepancies between this method and the actual RAP from the RHC, as it involved RHC data obtained by multiple different cardiologists without standardized ZRLs among them.

Out of the subjects in which this method proved incorrect, one had discordant values of right atrial pressure and right ventricular end diastolic pressure, which in the absence of mitral stenosis should be roughly equal. If RVEDP was used in place of RAP the qualitative prediction would be correct. One patient was incorrectly categorized as having JVD present due to the fact that the IJV was extremely dilated, yet completely collapsed with inspiration. In two patients who had severe tricuspid regurgitation, our technique underestimated RAP by 2-3 mmHg. Some RAP tracings had significant respiratory variation which may have altered the mean values, and some interventionalists asked the patient to exhale and hold their breath before each measurement while some did not. Also, given the large between-person variation in RAD, it is unclear if comparing this method which estimates the RAD in each patient is comparable to a ZRL that is a standard distance in every patient regardless of their individual right atrial depth. With any new technique, the proficiency of the operator improves as the study proceeds. This is evident by the fact that out of the last 25 patients, the qualitative prediction of RAP was correct 22 times.

This study was subject to several limitations. It was a single center study using a convenience sample of patients in need of right heart catheterization for any reason. It consisted of both outpatients and inpatients and no particular pathologies apart from congenital heart disease were excluded however this may have biased the results. The study sample was also skewed towards normal right atrial pressures. Since patients are often diuresed significantly prior to their RHC, only 10 of the 51 subjects had elevated pressures which may have affected the sensitivity and specificity analysis.

## Conclusion

In conclusion, this novel method for estimating JVP and RAP may offer a reliable way to rule out high right atrial pressure and could allow for more precise right atrial pressure estimation during physical examination.

## Data Availability

All data produced in the present study are available upon reasonable request to the authors

## LITERATURE CITED

1. Lewis T. Remarks on EARLY SIGNS OF CARDIAC FAILURE OF THE CONGESTIVE TYPE. Br Med J. 1930;1(3618):849–852.

2. McGee SR. Physical examination of venous pressure: A critical review. Am Heart J. 1998;136(1):10–18. doi:10.1016/S0002-8703(98)70175-9

3. Cook DJ. Clinical Assessment of Central Venous Pressure in the Critically Ill. Am J Med Sci. 1990;299(3):175–178. doi:10.1097/00000441-199003000-00006

4. Demeria DD, MacDougall A, Spurek M, et al. Comparison of Clinical Measurement of Jugular Venous Pressure Versus Measured Central Venous Pressure. Chest. 2004;126(4):747S. doi:10.1378/chest.126.4_MeetingAbstracts.747S

5. Davison R, Cannon R. Estimation of central venous pressure by examination of jugular veins. Am Heart J. 1974;87(3):279–282. doi:10.1016/0002-8703(74)90064-7

6. Connors AF, McCaffree DR, Gray BA. Evaluation of right-heart catheterization in the critically ill patient without acute myocardial infarction. N Engl J Med. 1983;308(5):263–267. doi:10.1056/NEJM198302033080508

7. Sinisalo J, Rapola J, Rossinen J, Kupari M. Simplifying the Estimation of Jugular Venous Pressure. Am J Cardiol. 2007;100(12):1779–1781. doi:10.1016/j.amjcard.2007.07.030

8. Brennan JM, Blair JE, Goonewardena S, et al. A Comparison by Medicine Residents of Physical Examination Versus Hand-Carried Ultrasound for Estimation of Right Atrial Pressure. Am J Cardiol. 2007;99(11):1614–1616. doi:10.1016/j.amjcard.2007.01.037

9. Bloomfield RA, Lauson HD, Cournand A, Breed ES, Richards DW. Recording of Right Heart Pressures in Normal Subjects and in Patients with Chronic Pulmonary Disease and Various Types of Cardio-Circulatory Disease. J Clin Invest. 1946;25(4):639–664. doi:10.1172/JCI101746

10. Seth R, Magner P, Matzinger F, Walraven CV. How Far Is the Sternal Angle from the Mid-right Atrium? J Gen Intern Med. 2002;17(11):861–865. doi:https://doi.org/10.1046/j.1525-1497.2002.20101.x

11. Kovacs G, Avian A, Olschewski A, Olschewski H. Zero reference level for right heart catheterisation. Eur Respir J. 2013;42(6):1586–1594. doi:10.1183/09031936.00050713

12. Deol GR, Collett N, Ashby A, Schmidt GA. Ultrasound Accurately Reflects the Jugular Venous Examination but Underestimates Central Venous Pressure. Chest. 2011;139(1):95–100. doi:10.1378/chest.10-1301

13. Simon Marc A., Schnatz Rick G., Romeo Jared D., Pacella John J. Bedside Ultrasound Assessment of Jugular Venous Compliance as a Potential Point-of-Care Method to Predict Acute Decompensated Heart Failure 30-Day Readmission. J Am Heart Assoc. 2018;7(15):e008184. doi:10.1161/JAHA.117.008184

14. Jang T, Aubin C, Naunheim R, Lewis LM, Kaji AH. Jugular venous distension on ultrasound: sensitivity and specificity for heart failure in patients with dyspnea. Am J Emerg Med. 2011;29(9):1198–1202. doi:10.1016/j.ajem.2010.07.017

15. Siva B, Hunt A, Boudville N. The sensitivity and specificity of ultrasound estimation of central venous pressure using the internal jugular vein. J Crit Care. 2012;27(3):315.e7-315.e11. doi:10.1016/j.jcrc.2011.09.008

16. New Method for Noninvasive Quantification of Central Venous Pressure by Ultrasound. doi:10.1161/CIRCIMAGING.114.003085

17. Lipton B. Estimation of central venous pressure by ultrasound of the internal jugular vein. Am J Emerg Med. 2000;18(4):432–434. doi:10.1053/ajem.2000.7335

18. De Vecchis R, Baldi C, Giandomenico G, Di Maio M, Giasi A, Cioppa C. Estimating Right Atrial Pressure Using Ultrasounds: An Old Issue Revisited With New Methods. J Clin Med Res. 2016;8(8):569–574. doi:10.14740/jocmr2617w

